# Breast density prediction from low and standard dose mammograms using deep learning: effect of image resolution and model training approach on prediction quality

**DOI:** 10.1101/2023.07.31.23293321

**Authors:** Steven Squires, Elaine Harkness, Alistair Mackenzie, D. Gareth Evans, Sacha J Howell, Susan M Astley

## Abstract

**Purpose:** To improve breast cancer risk prediction for young women, we have developed deep learning methods to estimate mammographic density from mammograms taken at approximately 1/10^th^ of the usual dose. We investigate the quality and reliability of the density scores produced on low dose mammograms focussing on how image resolution and levels of training affect the low dose predictions.

**Methods:** Deep learning models are developed and tested, with two feature extraction methods and an end-to-end trained method, on five different resolutions of 15,290 standard dose and simulated low dose mammograms with known labels. The models are further tested on a dataset with 296 matching standard and real low dose images allowing performance on the low dose images to be ascertained.

**Results:** Prediction quality on standard and simulated low dose images compared to labels is similar for all equivalent model training and image resolution versions. Increasing resolution results in improved performance of both feature extraction methods for standard and simulated low dose images, while the trained models show high performance across the resolutions. For the trained models the Spearman rank correlation coefficient between predictions of standard and low dose images at low resolution is 0.951 (0.937 to 0.960) and at the highest resolution 0.956 (0.942 to 0.965). If pairs of model predictions are averaged, similarity increases.

**Conclusions:** Deep learning mammographic density predictions on low dose mammograms are highly correlated with standard dose equivalents for feature extraction and end-to-end approaches across multiple image resolutions. Deep learning models can reliably make high quality mammographic density predictions on low dose mammograms.

## 1. Introduction

Early diagnosis of disease is an important factor in improving outcomes^1^ and better risk estimation can help to target scarce resources. Increasing personalisation of healthcare should enable early diagnosis or prevention of disease for those at higher risk while reducing negative effects for those at lower risk.^2^

An important risk factor for breast cancer is mammographic density^3^ where increased density is associated both with increased risk of developing breast cancer and reduced sensitivity of mammography for screening. While there are several methods of measuring mammographic density, visual assessment of mammography images by domain experts shows one of the strongest associations with future development of breast cancer.^4^ However, visual assessment is difficult to scale due to limitations in availability of medical expertise. Such limitations exist in wealthy countries but are even more pronounced in developing countries.^5^ Automated breast density models may be able to ameliorate the problem; machine learning methods can produce breast density estimates that correlate highly with radiologists’ visual assessment scores.^6^

The addition of mammographic density to risk models has been shown to improve performance^7^, but mammography is not recommended for young women which precludes the use of routine mammographic density assessment. Risk assessment mammography using a reduced radiation dose can potentially solve this problem. In this paper we refer to mammograms taken at the usual dose for a given women as ‘standard dose’ mammograms and those at a reduced dose (10% of standard) as ‘low dose’ mammograms.

Previous work has shown that there is a strong correlation between automated estimates of mammographic density for low and standard dose images.^8^ The quality of a prediction is a function of the information contained in the input together with the ability of the model to map to the output. The prediction quality compared to labels and the similarity between estimates on standard and low dose mammograms may depend on these two factors. The image quality is particularly impacted by the input image resolution which is often substantially reduced when training deep models while the quality of the deep model is dependent on, amongst other factors, the choice of feature extraction or fine-tuning methodologies. The image resolution and deep learning approach may cause variable conclusions based on model methodology rather than underlying science. The effect on standard and low dose images may be substantially different, with the potential for false conclusions to be drawn. For example, lower image resolution may obscure important features present in standard dose images that were never present in the low dose image, leading to falsely high correlations between the predictions on standard and low dose images. This would lead to a false belief that the low dose images could be used for risk prediction equivalently to standard dose images.

In this paper we test these ideas by taking one deep learning architecture and using two feature extraction versions and one fine-tuned version alongside images at five different resolutions. We train models on a dataset of standard dose image along with simulated low dose equivalents and test them on a separate dataset with both standard and low dose images. Our overall aim is to provide evidence as to whether mammographic density scores can reliably be produced on low dose mammograms, with a focus on the potential impact of image resolution and model training.

## 2. Material and methods

### 2.1 Data

In this study we utilise two datasets: one which consists of standard dose images and known mammographic density labels and the second consists of matched standard and low dose mammograms without reader annotated density scores.

#### 2.1.1 PROCAS dataset

To train and test our models we used a dataset from the *Predicting Risk of Cancer at Screening* (PROCAS)^9^ study, which consists of mammograms with associated reader visual analogue scale (VAS) density estimates. All PROCAS mammograms are from GE Senographe Essential (GE Healthcare, Buc, France) machines and have sizes of 3,062 by 2,394 or 2,294 by 1,914.

Two expert mammogram readers provided independent density estimates for both bilateral craniocaudal (CC) and mediolateral oblique (MLO) views. As the pair of readers was drawn on a pragmatic basis from a pool of 19, the VAS scores come from different pairs of readers. It has previously been shown that reader scores are variable^10,11^ but that the average of the two readers’ scores over all views has a strong association with breast cancer risk.^4^ The variability within and between readers may have some effect on model predictions.^12^ We have therefore selected a set of images scored by the same pair of readers to remove any variation due to inter-reader differences.

In total we have used 15,290 mammogram images from 4,051 women. We split the data so there are no images from the same woman in different partitions: 12,287 training images from 3,260 women; 1,533 validation images from 403 women; 1,470 test images from 388 women. Not all women in the dataset have four views, due to missing data.

All the images from the PROCAS study are standard dose so to enable us to train models to test on the ALDRAM low dose images we utilise a physics based modelling method that produces simulated low dose mammograms from standard dose images.^13,14^ The simulated low dose images are partitioned in the same manner as the standard dose images so that the separate models that we train on the standard dose and simulated low dose images alter only due to the nature of the images themselves along with the differences due to different training runs of any model.

The PROCAS dataset includes the mammograms of over 4,000 women and enables the models to be trained without substantial overfitting. The test set is sufficiently large that the uncertainties are small enough to be able to establish reliable conclusions. We can therefore be confident in the relationship between predictions on the simulated low dose and standard dose images. As we have expert reader labels (the VAS scores) we can also directly compare the predictions on the simulated low dose images to these labels with a high degree of confidence. However, we cannot be certain from these results that the models would provide the same conclusions on real low dose images.

#### 2.1.2 ALDRAM dataset

To assess the ability of models to accurately assess mammographic density in real low dose mammograms (as opposed to the simulated low dose images discussed previously) we used a dataset from the study *Automated Low Dose Risk Assessment Mammography* (ALDRAM),^8^ which consists of mammograms from 148 women. Each woman that consented to the study had a standard dose mammogram for CC and MLO views of the left and right breasts. While under compression for the views of the right breast the milliampere-seconds (mAs) were reduced by 90%, or to a minimum of 3mAs, whichever was higher, with peak kilovoltage (kVp) and filters unchanged and another image taken (the low dose mammogram). There are therefore six mammographic images for each of the 148 women: CC and MLO views of both breasts with a standard dose and RCC and RMLO views with a low dose. All ALDRAM images were taken using a GE Senographe Essential machine (GE Healthcare, Buc, France). Images were either 3,062 by 2,394 or 2,294 by 1,914 pixels in size. In this study we only use unprocessed (for processing) images, and no visual assessment of density is available.

The ALDRAM dataset is kept as an independent test set because it is too small to train deep learning models on whilst also being able to validate the results but also because keeping it independent means we can be confident we are not overfitting our models to this dataset. The combination of low and standard dose images is not available elsewhere so these result on this test set are a highly valuable resource to enable judgement of the capacity of deep learning models to make mammographic density predictions on low dose images. The fact that the low and standard dose images are equivalent in all other ways means we can isolate the differences in performance to those differences in the dose.

### 2.2 Experimental approach

The combination of the large PROCAS dataset with associated VAS scores and the generated simulated low dose images means we can form a large comparison study between the quality of predictions on simulated low dose images compared to standard dose images. The addition of the real low dose images with equivalent standard dose images in the ALDRAM datasets means we can produce evidence of the capacity of deep learning models to estimate mammographic density scores, and hence cancer risk predictions, on low dose images.

We utilise a ResNet^15^ model to make predictions, the details of which we will discuss in Section 2.3. We train and run the models on five resolutions: 80 by 64, 160 by 128, 320 by 256, 640 by 512, and 1,280 by 1,024, which we designate as resolutions 1 to 5 respectively. We train our models on standard dose images from the PROCAS dataset as well as the equivalent simulated low dose images. The trained models are then used for inference on the test sets from PROCAS and ALDRAM.

The ResNet model is a popular model which produces accurate predictions on multiple tasks. These include previous models trained to make mammographic predictions which produce state-of-the-art performance.^12^ Our questions relate to the reliability of deep learning mammographic density predictions on low dose images especially considering the image resolution and training of the models rather than trying to find a model which maximises performance.

The input images are unprocessed (for processing) mammograms either 3,062 by 2,394 or 2,294 by 1,914 pixels in size. These are pre-processed to reduce the resolution in a way that maintains the aspect ratio of the original images. Smaller images are padded with background pixels to increase the height to 2,995 pixels and the width 2,394 pixels and larger images are cropped to the same size. Cubic interpolation is used to generate images of the five different resolutions. Pixel values are then clipped to 75% of the maximum, the pixel values inverted, histogram equalisation applied and the values normalised so that the final pixel values were between 0 and 1. Before running through the ResNet models the greyscale images are copied across the three input channels and normalised to have means of [0.485,0.456,0.406] and standard deviations of [0.229,0.224,0.225] to match the ImageNet^16^ images the ResNet models were trained on.

### 2.3 Models

To investigate the importance of model quality we use three different deep learning models all with a ResNet-18^15^ core. Two of these are based on feature extraction with a linear regression model used on the extracted feature vector while the third is trained end-to-end.

#### 2.3.1 Feature extraction models

The first feature extraction model, which we refer to as *untrained*, has randomised weights and the second, which we refer to *pretrained*, as has weights taken from its training on ImageNet^16^. The pretrained model would be expected to show superior performance than the untrained version.

To train these versions of ResNet we truncate the model, removing the final standard connected layer, which results in a *p = 512* dimensional output, *v*i. We concatenate the *n*tr training images into a matrix, 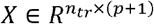, with the additional of a bias value to the 512-dimensional vector. We then use the pseudo-inverse to train a linear regression to find the weight vector, *w*, which minimises the mean squared error, selecting the *L*_2_ regularisation level with the validation set. This approach has shown good performance on both the ALDRAM^8^ and PROCAS^17^ datasets previously.

This procedure is carried out on the PROCAS standard dose and simulated low dose images to find the weight vectors for each set of images for each resolution. The appropriate weights are then applied to the feature vectors extracted from the two test sets.

#### 2.3.2 End-to-end trained models

For the end-to-end trained models (named *trained)* we train on both CC and MLO images. The models are initialised with pre-trained weights found via training on ImageNet. We remove the final fully connected layer of the ResNet and replace it with a single linear layer connected to one output neuron. This end-to-end model is a fine-tuned version of the *pretrained* model. For data augmentation we utilise random left-right flips and the addition of a small amount of Gaussian random noise (centred at zero and with a scale of 0.01). We also train models without data augmentation. The Adam^18^ optimiser was used for all trained models with only the learning rate changed from defaults.

We use mean squared error (MSE) as an objective function. On each epoch we assess the MSE on the validation set and save the model parameters if the validation MSE is lower than any previous epoch.

We performed multiple training runs on all image resolution varying the learning rates and use of data augmentation. We selected our final models by comparing the Spearman rank correlation coefficient between the predictions made by the models on the validation set of the PROCAS (standard dose or simulated low dose) dataset and the average density as assessed by the pair of readers. We trained all our end-to-end models on a single Nvidia V100 GPU (Nvidia Corporation, California, USA) with 16GB GPU RAM using Pytorch.^19^

## 3. Results

In Table 1 we show Spearman’s rank correlation coefficients and the root mean squared error (RMSE) of our model predictions for the five image resolutions compared to the average reader density, using standard and simulated low dose PROCAS images. The purpose of this table is to show the importance of the image resolution and model quality with respect to the labels for the standard and simulated low dose images. Confidence intervals are generated using bootstrapping and reported at the 95% level.

**Table 1.**
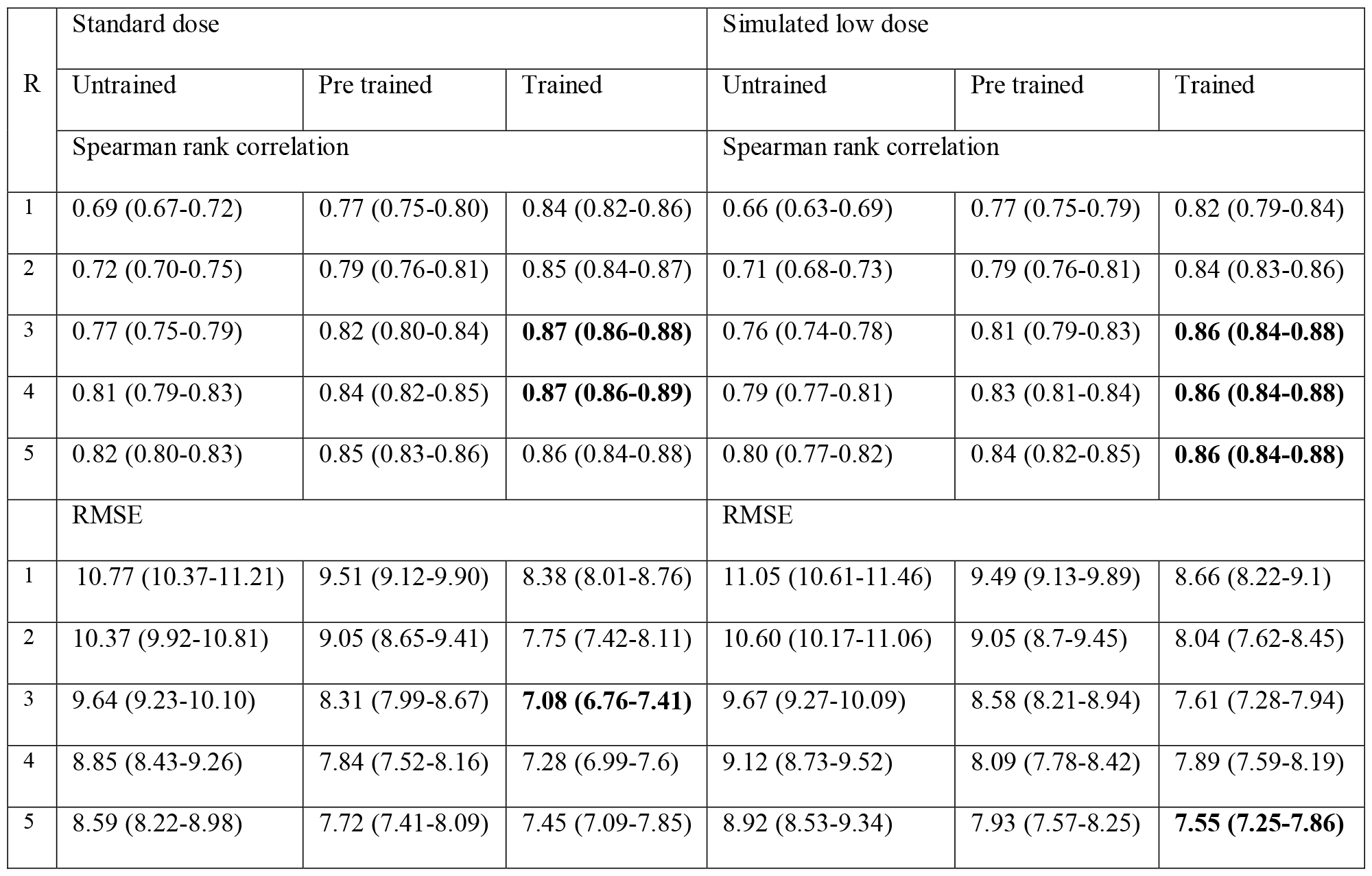
Spearman’s rank correlation coefficient (top set of rows) and RMSE (bottom set of rows) for the predictions made by the models trained and tested on PROCAS images, standard and simulated low dose, produced at the five different resolutions (denoted as R) compared to the labels provided by averaging scores provided by radiologists. The best performing models are shown in bold separately for the two metrics and the standard and simulated low dose images.

To demonstrate how the similarity varies with resolution between standard and low dose predictions we show, in Figure 1, Spearman’s rank correlation coefficient scores between the model predictions of the PROCAS standard dose and simulated low dose images test sets. The untrained model shows a rise in consistency between low dose and standard dose before the similarity falls back again at resolution 5. The pretrained model shows an increase in rank correlation coefficient as the resolution increases followed by a plateau at the higher resolutions. The trained model generally shows an increase in prediction similarity as the resolution is increased.

**Figure 1.**
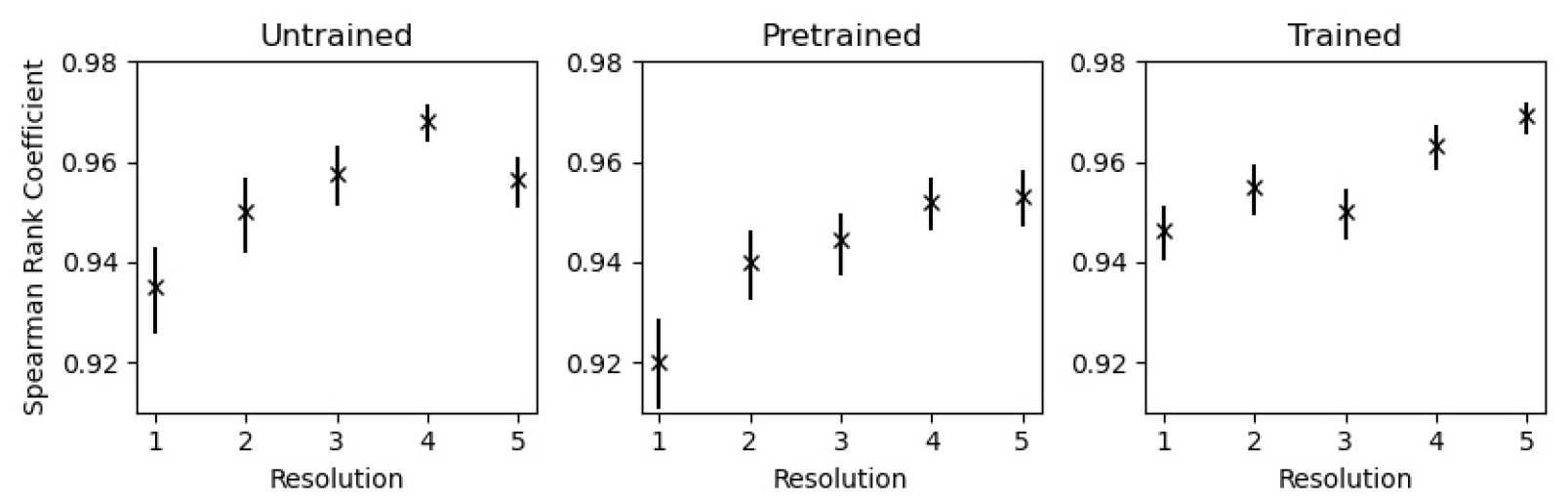
Spearman’s rank correlation coefficient between predictions on the PROCAS standard dose and simulated low dose images at the five different resolutions for the three different model variants.

In Figure 2 we show a comparison between the model predictions of the ALDRAM standard dose and low dose images at the different image resolutions. The uncertainty is higher than for the simulated low dose images (in Figure 1) due to the smaller amount of data.

**Figure 2.**
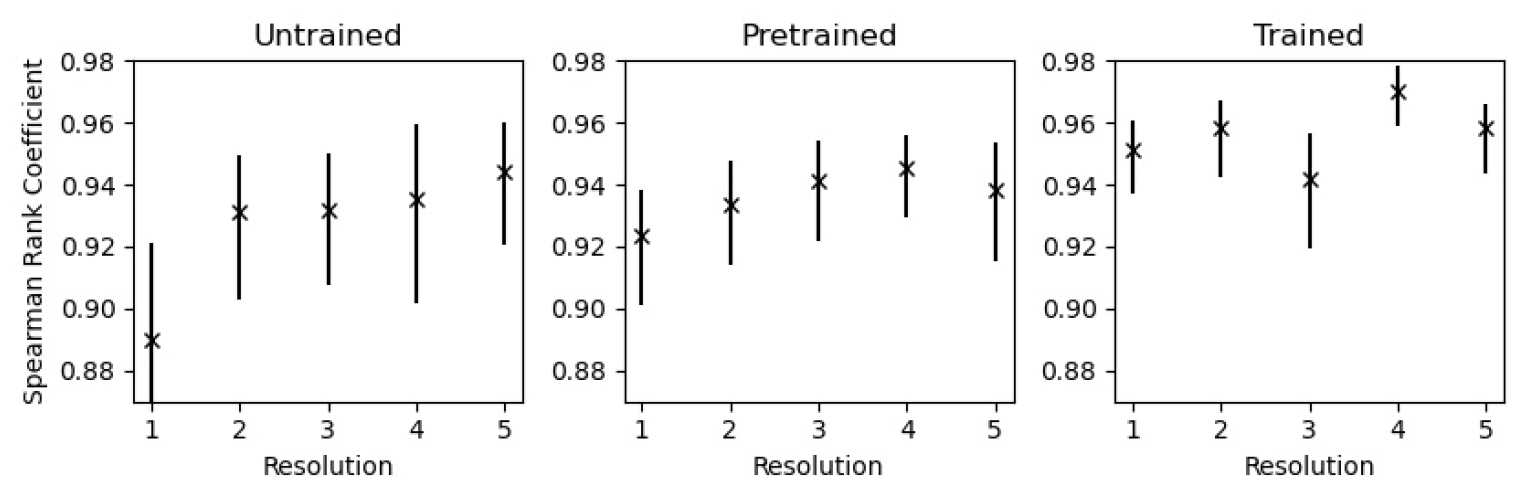
Spearman’s rank correlation coefficients between model predictions on the ALDRAM standard dose and low dose for the three model variants at the different image resolutions.

To attempt to reduce some of the variation due to stochasticity in training we take the average predictions of the two best performing trained models for the same resolution. We then plot the averaged low dose predictions against the averaged standard dose predictions for the ALDRAM data. These results are shown in Figure 3 with the Spearman rank correlation coefficient noted in the top left corner. The correlation between the low dose and standard dose predictions is higher than when we did not average.

**Figure 3.**
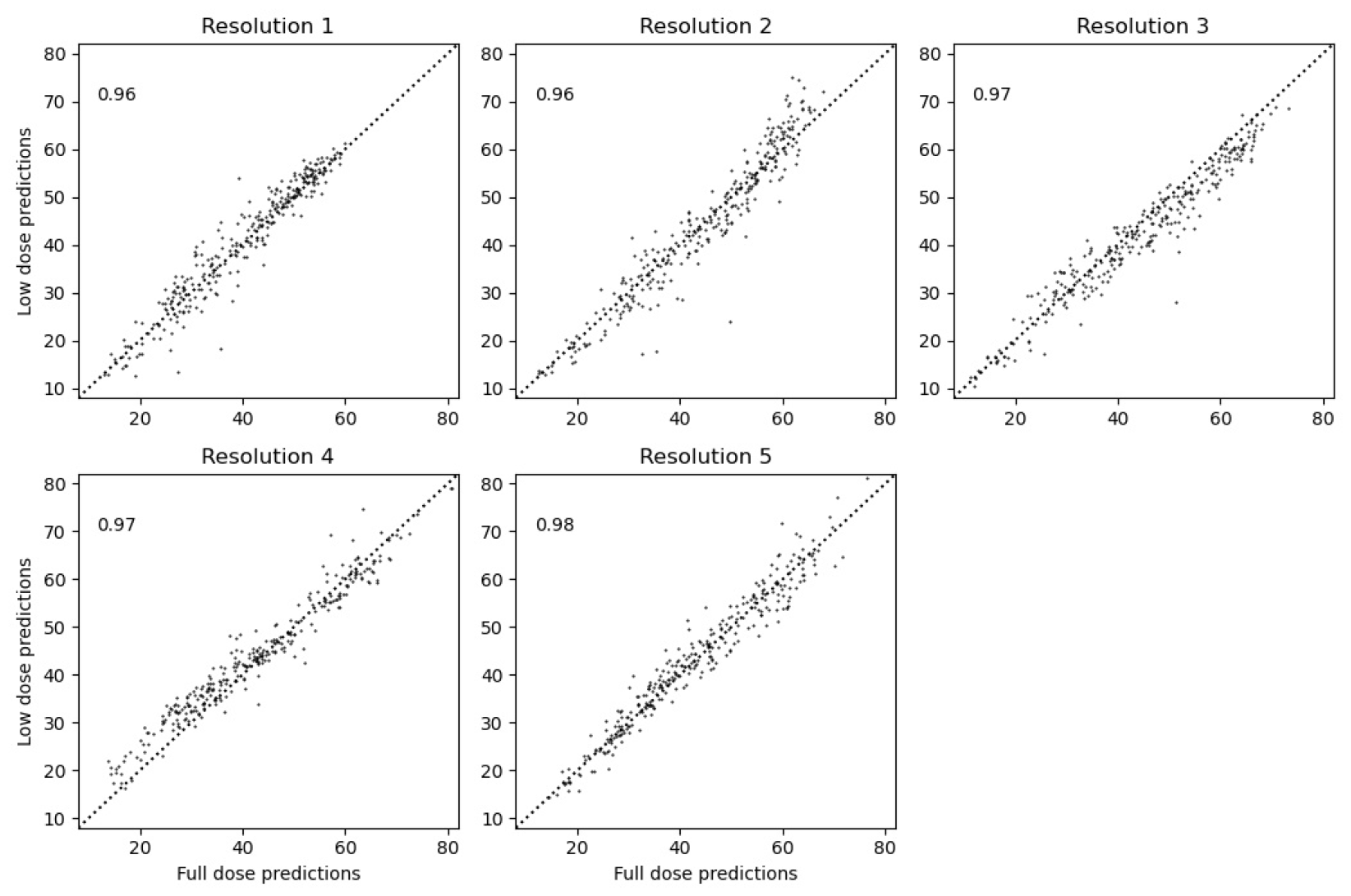
Plots of ALDRAM predictions between standard and low dose images when averaged across the two best performing (defined as the lowest Spearman rank correlation on the PROCAS validation set) trained models for both standard dose and simulated low dose trained models. Spearman’s rank correlation is shown in the top left corner.

## 4 Discussion

The aim of this study was to investigate how two important factors in deep learning models, image resolution and level of training, affect mammographic density estimates on both standard and low dose images. Of particular importance is how the differences in model methodology might affect conclusions drawn about the comparisons of predictions on the standard and low dose images.

The trained models perform better than untrained or pretrained models including showing good performance at low image resolutions, where the level of information content might be thought to be too low. They show no improvement beyond resolution 3 (320 by 256) which would need to be investigated further but is likely due to some combination of quantity of data and the effect of the label variability on both training and assessment of the quality of the models which has previously been shown to limit apparent model performance.^17^

Increasing the image resolution has a substantial impact on the untrained and pretrained models with the pretrained model showing performance almost equal to that of the trained model at the highest resolution. This effect implies, contrary to the evidence of the trained model, that increasing resolution improves model performance. A possible solution is that that label variability is a constraint on all the models, that both image resolution and improved model training would result in improved performance, but the label variability limits this.

The prediction quality compared to labels is similar for both standard and simulated low dose images. The small differences in performance are mostly not statistically significant. The density labels are known to show a strong relationship to cancer risk^4^ which has also been shown with the model predictions on standard dose images^6,17^. This implies the predictions on the low dose images will show equivalent capacity to estimate cancer risk.

The similarity between predictions on standard and simulated low dose images does not fall with image resolution, which would be expected if resolution reduction was obscuring features in the standard dose images that were never available in the simulated low dose images. If there is a change it is more likely that the similarity improves with resolution.

When we test the models on the ALDRAM data of standard and low dose images we see high levels of correlation between predictions. If we reduce general model variability by averaging over pairs of models the similarity increases further up to a Spearman rank correlation coefficient of 0.98 at the highest resolution, which shows the low dose predictions are highly correlated with the standard dose predictions.

Overall, the evidence is that deep learning models can make predictions of mammographic density on low dose images that are very similar to those on standard dose images. It is advantageous to train models end-to-end but if a large enough dataset is not available using feature extraction with linear regression works well. As risk estimates can be used from standard dose images these results provide strong evidence that risk estimates of comparable quality can be extracted from the low dose images.

## 5. Conclusions

Low dose mammography has the potential to provide breast cancer risk assessment, via mammographic density prediction, for women who are not recommended to have standard dose mammograms taken. We have shown that deep learning models can make estimates of mammographic density that correlate highly with standard dose equivalents. Fine-tuned deep models perform better than feature extraction-based models with linear regression but both perform well.

Image resolution reductions could conceal relevant features in the standard dose images that would not have been available in the low dose equivalents. This would have led to higher-than-expected correlations between standard dose and low dose model predictions, however we show that this is not that case and that the reduction in image resolution does not confound our conclusion that deep learning models can make accurate mammographic density predictions on low dose images.

Overall, we have demonstrated that the combination of low dose mammography with deep learning models is able to produce density-based risk estimates comparable to those of standard dose images. Therefore, schemes to use low dose mammography should be able to rely on models producing reliable cancer risk estimates. There are small (and mostly not statistically significant) differences in performance on the low dose images compared to standard dose which should be investigated further to either explain the differences or find methods to improve the performance on the low dose images.

## Data Availability

Data is not currently available outside the University of Manchester

